# A high centrifugal force-enhanced Ziehl-Neelsen method for improved detection of *Mycobacterium tuberculosis*

**DOI:** 10.64898/2026.01.21.26344580

**Authors:** Godlove T Chaula, Lucy Namkinga, Wilber Sabiiti, Nyanda E Ntiningya, Bariki Mtafya, Ally Mahadhy

**Affiliations:** National Institute for Medical Research (NIMR), Mbeya Medical Research Centre (NIMR-MMRC), Hospital Hill Road 1, Mbeya, Tanzania; University of Dar es Salaam, College of Natural and Applied Sciences, Department of Molecular Biology and Biotechnology, P. O. Box 35179, Dar es salaam, Tanzania; University of Dar es Salaam, Mbeya Collage of Health and Allied Sciences, Department of Biochemistry & Pharmacology, P. O. Box 608, Mbeya; University of St Andrews, School of Medicine, St. Andrews, United Kingdom

**Keywords:** Relative centrifugal force, *Mycobacterium tuberculosis*, ZN-smear microscopy. Centrifugation, MTB Diagnosis

## Abstract

Ziehl–Neelsen (ZN) smear microscopy remains central to tuberculosis (TB) diagnosis and treatment monitoring, yet its sensitivity is limited by incomplete recovery of *Mycobacterium tuberculosis* during pre-analytical processing. This study evaluated whether modifying centrifugation force and duration improves bacillary recovery and ZN smear performance. Laboratory experiments were conducted using *M. tuberculosis* H37Rv and pulmonary TB sputum samples. Following NALC–NaOH decontamination, samples were centrifuged at 2,000, 3,000, or 6,000 × g for 40 min, and the effect of centrifugation time at 3,000 × g was assessed by comparing 20 and 40 min using the same specimens. ZN smear grading and positivity were evaluated in triplicate and compared statistically, with significance set at *P* < 0.05. In H37Rv suspensions, smear grading increased with higher centrifugal force, while smear positivity plateaued at 3,000 × g. In contrast, clinical sputum samples showed progressive increases in both smear grading and positivity with increasing centrifugal force, with statistically significant improvements in positivity (*p* = 0.0097). Extending centrifugation time at 3,000 × g did not change smear positivity in laboratory suspensions or clinical sputum (*P* = 0.30). These findings indicate that standard centrifugation conditions are sufficient for laboratory strains, whereas higher centrifugal forces enhance bacillary recovery from clinical sputum, improving ZN smear sensitivity. Optimizing relative centrifugal force during pre-analytical processing may therefore reduce false-negative results and strengthen the diagnostic and treatment-monitoring performance of ZN smear microscopy in routine TB laboratories.

## 1. Introduction

Tuberculosis (TB) remains a major public health challenge, claiming over 1 million lives annually out of an estimated 10 million people who contract the disease each year [1]. Controlling TB requires not only effective diagnostics and treatment but also innovative tools to monitor treatment response. Despite the availability of effective treatment regimens, TB elimination has yet to be achieved, primarily due to significant diagnostic gaps. In 2022, approximately 30% of individuals with TB were not diagnosed and, consequently, did not receive treatment [2]. Moreover, 37% of diagnosed cases were based solely on clinical assessment, which often lacks sufficient specificity and sensitivity [3] – [4]. These persistent diagnostic shortcomings underscore the urgent need to evaluate current screening strategies and optimize the use of existing diagnostic tools to enhance TB case detection and bridge the diagnostic gap.

The prolonged duration of tuberculosis (TB) treatment, along with the use of multiple toxic drug regimens, not only incurs high costs but also poses serious safety concerns for patients [5]. The situation is further complicated by the rise of drug-resistant TB (DR-TB), which demands even more complex, toxic, and extended treatment compared to drug-susceptible TB (DS-TB). This underscores the critical need for effective tools to monitor treatment response [6]. Such tools would enable early detection of patients who are not responding well to therapy or are at risk of relapse or treatment failure [7] – [8]. Currently, Ziehl-Neelsen (ZN) smear microscopy is the standard diagnostic approach for TB [9]. Whereas, the combination of N-Acetyl-L-Cysteine and Sodium Hydroxide (NALC-NaOH) followed by standard centrifugation at 3000 х g for 15 – 20 min is the most widely used method for pre-analytical sample processing prior to ZN smear microscopy [10] – [11]. However, this approach suffers from several technical drawbacks, including low sensitivity due poor recovery during centrifugation [12] – [13].

Given the low specific gravity of *Mycobacterium tuberculosis* (ranging from 1.07 to 0.79), a high RCF is necessary to effectively concentrate the bacilli into the pellet [14]. However, previous research has shown that the current standard centrifugation settings of 3000 – 3500 × g for 20 minutes lead to over 90% unrecovered mycobacterial cells from the pellet [15]. Therefore, there is a pressing need to re-evaluate the current standard centrifugation protocol 3000 × g for 20 minutes to enhance the recovery of *M. tuberculosis* for the ZN smear microscopy. In this context, we examined the effects of increasing centrifugal force by varying the centrifugal speeds at fixed duration for the recovery of *M. tuberculosis* cells for of ZN smear microscopy.

## 2. Materials and Methods

### 2.1 Study samples

In this study, *Mycobacterium tuberculosis* H37Rv (ATCC 27294) laboratory strain and clinical pulmonary tuberculosis sputum samples were used to assess the impact of relative centrifugal force at a fixed duration on the recovery of *M. tuberculosis*, with subsequent Ziehl–Neelsen (ZN) smear microscopy. The clinical sputum samples were obtained from adult participants aged 18–65 years who were recruited at Rungwe District Hospital, Mbeya, Tanzania, between 26/05/2021 and 26/05/2022, as part of the TB-MBLA translational study. Written informed consent was obtained from all participants prior to enrolment; for participants unable to read or write, witnessed verbal consent was obtained in accordance with approved ethical procedures.

### 2.2 Ziehl–Neelsen Smear Grading and Positivity

The effect of relative centrifugal force (RCF) on the recovery of *Mycobacterium tuberculosis* from samples was assessed using Ziehl–Neelsen (ZN) smear microscopy, with both smear grading and positivity determined as previously described [14]. Following acid-fast staining, bacilli were observed and counted under a microscope. Smears were graded according to the number of acid-fast bacilli (AFB) observed: if no AFB were detected in 100 fields, the result was considered negative; 1–9 AFB in 100 fields was classified as scanty; 10–99 AFB in 100 fields was graded as 1+; 1–10 AFB per field in at least 50 fields was graded as 2+; and more than 10 AFB per field in at least 20 fields was graded as 3+. ZN smear positivity (%) was then calculated using the formula [16]:

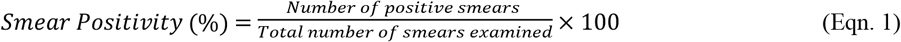

### 2.3 Effect of Centrifugal Force on Recovery of *M. tuberculosis* H37Rv

*In vitro* experiments were performed using a 2 weeks old culture of *M. tuberculosis*, H37Rv strain (ATCC 27294) grown in Lowenstein Jensen (LJ) medium. The *M. tuberculosis* culture was adjusted to 0.5 McFarland (approximately 1.0 x10^8^ CFU/ml) and serially diluted in Middlebrook 7H9 broth to 100 CFU/ml). The resulting dilutions were decontaminated using 1 % N-Acetyl-Cysteine and 2% Sodium hydroxide (NALC-NaOH) for 20 minutes. The pellets were recovered by centrifugation at 2000 × g, 3000 × g, and 6000 × g for 40 minutes at room temperature [17]. The pellets were suspended in 2 mL of PBS and 50 µL (× 3) of it was used to prepare smears. Results was confirmed by ZN smear microscopic examination and recorded as per WHO guidelines [16].

### 2.4 Effect of Centrifugal Force on Recovery of *M. tuberculosis* in Clinical Sputum Samples

Pulmonary tuberculosis (TB) patient’s specimens were retrieved from a -80°C freezer. The samples were transferred to a -20°C freezer overnight and thawed at 2-8°C at room temperature. The samples were homogenized using a sterile magnetic stirrer for 20 min at room temperature. A 2 mL of pooled sputum was decontaminated with NALC/NaOH for 20 min. The pellets were recovered by centrifugation at 2000 × g, 3000 × g, and 6000 × g for 40 minutes at room temperature [17]. The recovered pellets were then suspended in 2 mL of PBS, and 50 µL (× 3) of it was used to prepare smears. Results were confirmed by ZN smear microscopic examination and recorded according to WHO guidelines [16].

### 2.5 Quality Control and Replicates

For the H37Rv laboratory strain, each relative centrifugal force (RCF) condition was tested in five independent replicates. For clinical sputum samples, ten specimens were processed, and each RCF condition was tested in triplicate for every specimen. A control setting of 3,000 × g for 20 min was included in all experiments to account for any non-specific effects. Additionally, all centrifugation steps were performed at 4 °C, and both the centrifuge and microscope were calibrated prior to each experiment to ensure optimal performance and reproducibility.

### 2.6 Data Analysis

Data were recorded in Microsoft Excel and analyzed using GraphPad Prism version 7.04 (GraphPad Software, La Jolla, CA, USA). ZN smear positivity was expressed as percentages of positive samples, while smear grades were summarized as medians. Differences in smear outcomes across centrifugal forces (2,000 × g, 3,000 × g, and 6,000 × g for 40 min) were assessed using the Kruskal–Wallis test. Comparisons involving the same specimens processed under two conditions (e.g., 3,000 × g for 20 min vs. 3,000 × g for 40 min) were performed using the Wilcoxon signed-rank test. All tests were two-sided, and *P* < 0.05 was considered statistically significant.

## 3. Results and Discussion

### 3.1 Recovery of *M. tuberculosis* H37Rv at Different RCF (ZN Microscopy)

M. tuberculosis H37Rv suspensions were centrifuged at 2,000, 3,000, and 6,000 × g, and ZN smear grading and positivity were assessed (Fig 1). Smear grading increased progressively with higher RCF, indicating that stronger centrifugal forces concentrated more bacilli into the sediment. In contrast, positivity followed a different trend: 40% ± 3% of aliquots were positive at 2,000 × g, compared with 60% ± 5% and 60% ± 2.6% at 3,000 × g and 6,000 × g, respectively. This difference was not statistically significant (*p* = 0.3), likely due to the small number of replicates (n = 5 per RCF) and the inherent variability in bacilli distribution within each sample, which reduced statistical power. These results suggest that increasing the RCF from 2,000 × g to 3,000 × g improved bacillus detection, but no further gain was achieved at 6,000 × g, indicating a plateau effect.

**Fig 1.**
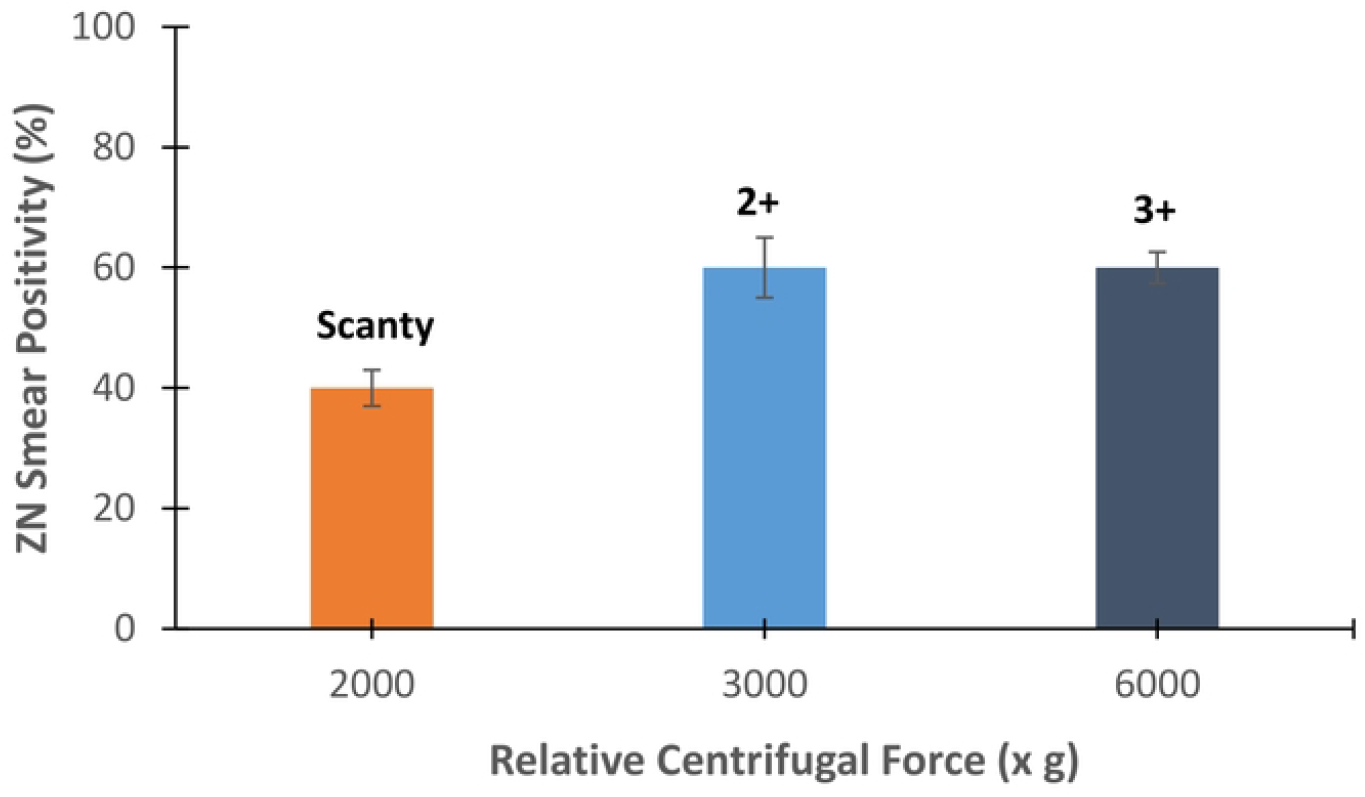
Median ZN smear grade and smear positivity (%) of *M. tuberculosis* H37Rv at different relative centrifugal forces (RCFs). Positivity represents the percentage of positive smears at each RCF.

ZN smear positivity (%) of *M. tuberculosis* H37Rv at different RCFs. Grading increased with higher RCFs and positivity increased from 2,000 × g to 3,000 × g and plateaued at 6,000 × g. To further explore the influence of spin time, a control experiment was performed at 3,000 × g for 20 min. This shorter centrifugation yielded a smear grading of 2+ and positivity of 55% ± 4.5%, slightly lower than the 60% ± 5% observed at 3,000 × g for 40 min. The paired difference did not reach statistical significance (*p* = 0.15), indicating that while reducing spin time modestly decreases sedimentation efficiency, the majority of bacilli are recovered within the first 20 minutes at this RCF.

The variability in positivity across aliquots at the same RCF can be explained by heterogeneity in bacilli distribution within the suspension. At lower RCF (2,000 × g), sedimentation efficiency is limited, leading to uneven recovery and some aliquots lacking sufficient bacilli to cross the detection threshold. At 3,000 × g, sedimentation is more effective, reducing this variability and yielding higher positivity. Beyond this point, however, the proportion of positive aliquots does not increase further, likely because most bacilli capable of being pelleted have already been recovered.

Interestingly, although positivity plateaued at 3,000 × g, smear grading continued to rise at 6,000 × g, reflecting a greater bacillary load per smear field. This indicates that higher RCFs enhance semi-quantitative assessment of bacterial burden even when the proportion of positive samples remains unchanged.

In practical terms, an RCF of 3,000 × g appears optimal for the *M. tuberculosis* H37Rv laboratory strain, as it maximizes positivity and improves sample consistency. Higher RCFs such as 6,000 × g may still provide value by increasing smear grading, further concentrating bacilli per field. These findings emphasize the importance of considering both centrifugal force and sample heterogeneity when optimizing protocols for ZN smear microscopy.

### 3.2 Recovery of *M. tuberculosis* from Sputum at Different RCF (ZN Microscopy)

The efficiency of *M. tuberculosis* recovery from clinical sputum samples was also evaluated at different RCFs (2,000, 3,000, and 6,000 × g) using ZN smear grading and positivity (Fig 2).

**Fig 2.**
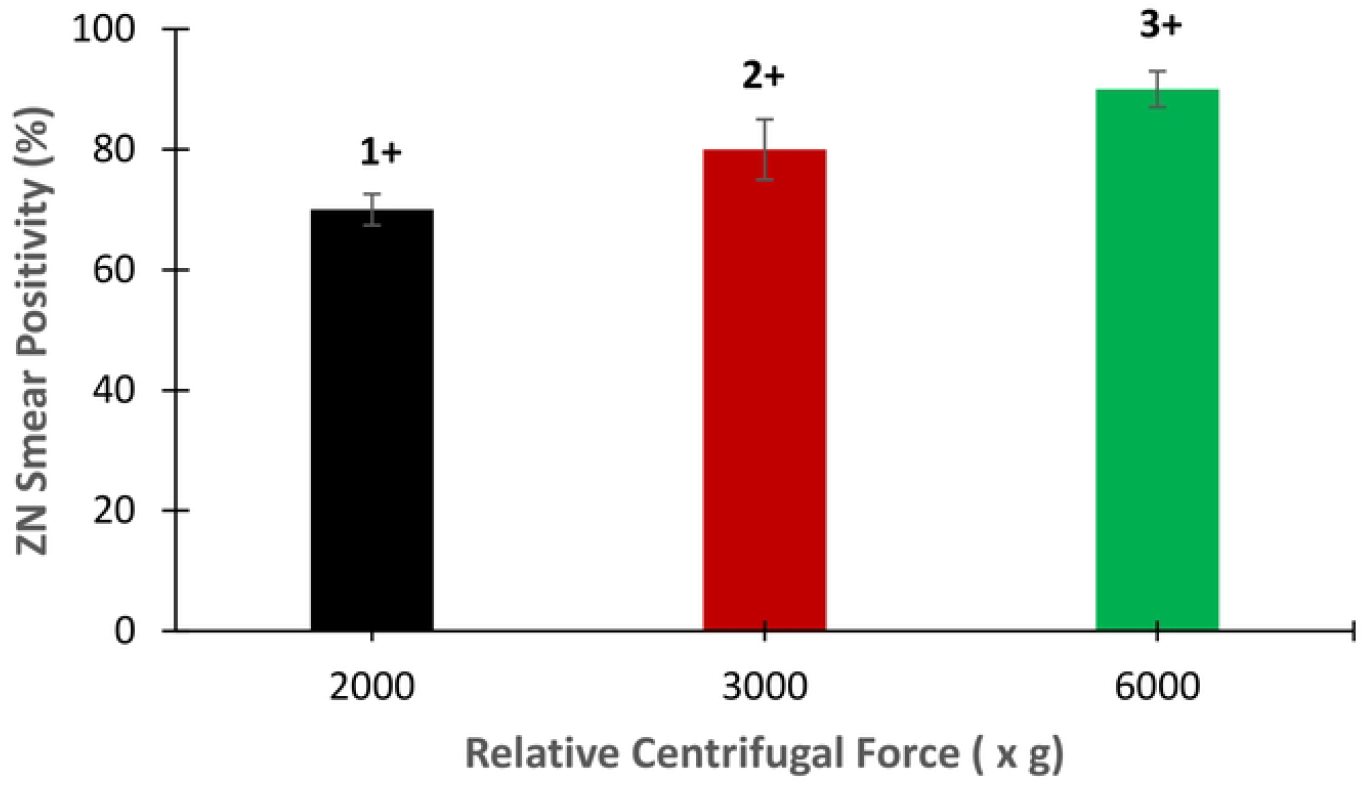
Median ZN smear grade and smear positivity (%) of *M. tuberculosis* in clinical sputum at different relative centrifugal forces (RCFs). Positivity represents the percentage of positive smears at each RCF.

In contrast to the H37Rv laboratory strain, smear grading and positivity of clinical sputum increased with higher RCFs. At 2,000 × g, incomplete sedimentation likely limited bacilli recovery, resulting in lower grading and fewer positive samples. Increasing the force to 3,000 × g improved both parameters, and further increasing to 6,000 × g continued to enhance recovery, indicating that higher centrifugal forces more effectively concentrated bacilli from the heterogeneous sputum matrix. Positivity differed significantly among RCFs (*p* = 0.0097), confirming that higher centrifugal force improved bacillus detection. These findings suggest that centrifugation efficiency plays a more critical role in clinical specimens, where bacterial load and distribution are more variable than in standardized laboratory suspensions. They are also in agreement with previously published data [14] and reflect the complex nature of clinical sputum, in which mucus, host cells, and debris can hinder bacilli sedimentation, necessitating greater centrifugal force for efficient recovery.

To examine the effect of centrifugation duration, a control experiment was conducted at 3,000 × g for 20 min. This shorter spin resulted in a smear grading of 1+ and positivity of 75% ± 3%, slightly lower than the 80% ± 5% observed at 3,000 × g for 40 min; however, the difference was not statistically significant (*p* = 0.12). These results indicate that while most bacilli are pelleted within the first 20 minutes, extending the spin time modestly improves recovery, particularly in clinical sputum where bacilli are embedded in mucus and cellular debris.

The progressive increase in both grading and positivity across RCFs in sputum samples highlights the importance of optimizing centrifugation conditions for clinical diagnostics. Unlike the laboratory strain, where positivity plateaued at 3,000 × g, clinical samples continued to benefit from higher centrifugal forces. This underscores that optimal centrifugation parameters may differ between standardized laboratory suspensions and patient-derived specimens.

From a practical perspective, applying higher RCFs, such as 6,000 × g, in routine diagnostic protocols may enhance the sensitivity of ZN smear microscopy for sputum samples, particularly in cases with low or uneven bacillary loads. While moderate RCFs may suffice for laboratory strains, clinical specimens require stronger centrifugal forces to maximize bacilli recovery and improve diagnostic yield. Mycobacterial species have low specific gravity, necessitating higher centrifugation speeds and longer durations for optimal pellet recovery [10]. Processing samples at lower centrifugation speeds may result in the loss of some *M. tuberculosis* bacilli, potentially turning downstream test results from positive to negative. This could lead to misdiagnosis, inaccurate treatment monitoring, and failure to identify non-responding patients by the second month of TB therapy, potentially contributing to relapse or the development of drug-resistant TB, which is costlier and requires longer treatment [18] – [19].

Although WHO-recommended, highly sensitive DNA-based molecular tools such as Xpert MTB/RIF are beneficial for TB diagnosis, treatment monitoring still relies heavily on culture and less sensitive sputum smear microscopy [20]. Therefore, optimizing sample processing methods, including adjustments to relative centrifugal force and duration, could enhance recovery of low bacillary loads, thereby improving the accuracy of TB diagnosis, treatment evaluation, and monitoring.

## 4. Conclusion

This study demonstrates that the recovery of Mycobacterium tuberculosis in ZN smear microscopy is strongly influenced by the relative centrifugal force applied during pre-analytical processing. For the laboratory H37Rv strain, an RCF of 3,000 × g was sufficient to maximize positivity, with higher forces primarily enhancing smear grading. In contrast, clinical sputum samples benefited from progressively higher RCFs, with 6,000 × g improving both grading and positivity, highlighting the greater variability and complexity of patient-derived specimens. These findings underscore the critical role of optimizing centrifugation parameters to improve bacilli recovery, particularly from heterogeneous clinical samples. Implementing higher centrifugal forces in routine diagnostic protocols may enhance the sensitivity of ZN smear microscopy, reduce the risk of false-negative results, and support more accurate treatment monitoring, ultimately contributing to better TB management and control.

## Data Availability

All relevant data are within the manuscript

## Acknowledgment

We sincerely thank the research support team from the Infection Group of the University of St Andrews, United Kingdom, and the laboratory team from the National Institute for Medical Research (NIMR), Mbeya Medical Research Centre, Tanzania, respectively, for their invaluable technical support. We also acknowledge the NIMR-Mbeya Medical Research Centre for providing the research infrastructure and resources necessary to conduct this study, and we are grateful to the patients who generously provided samples for the laboratory experiments. This work was supported by the European and Developing Countries Clinical Trials Partnership (EDCTP) through the PanaCEA consortium and forms part of the MSc thesis of Mr. Godlove Chaula at the University of Dar es Salaam.

## Ethical statement

This work was nested in the TB-MBLA translational study in routine healthcare practice conducted in Mbeya, Tanzania. The study was approved by the Mbeya Medical Research and Ethics Committee (SZECEC-2439/R. A/V.1/83) and Medical Ethics and Research Coordinating Committee of the National Institute for Medical Research (NIMR/HQ/R.8a/Vol.IX/3687). The patients offered a written consent or witnessed verbal consent for those who could not write or read to authorize the use of samples for research purposes and validation of novel TB diagnostics.

## Authors’ contributions

**GC, LN, BM** and **AM** designed the study; **GC** performed the experiments; **GC, LN, BM** and **AM** analyzed the data; **GC and AM** wrote the first draft of the manuscript; **BM, WS** and **NN** solicited research funds; All authors (**GC, LN, WS, NN, BM** and **AM**) reviewed and approved the manuscript before submission.

